# Covid-19 vaccination intentions among Canadian parents of 9-12 year old children: results from the All Our Families longitudinal cohort

**DOI:** 10.1101/2020.11.24.20237834

**Authors:** Erin Hetherington, Sarah A Edwards, Shannon E MacDonald, Nicole Racine, Sheri Madigan, Sheila McDonald, Suzanne Tough

**Affiliations:** Department of Obstetrics & Gynaecology, Cumming School of Medicine, University of Calgary; Department of Epidemiology, Biostatistics & Occupational Health, McGill University; Department of Community Health Sciences, Cumming School of Medicine, University of Calgary; Department of Pediatrics, Cumming School of Medicine, University of Calgary; Faculty of Nursing, University of Alberta; Department of Psychology, University of Calgary; Alberta Children’s Hospital Research Institute

## Abstract

**Background:** Acceptance of a COVID-19 vaccine is critical to achieving high levels of immunization. The objective of this study is to understand factors associated with COVID-19 vaccine intentions among parents and explore reasons underlying decision making.

**Methods:** Participants from a longitudinal cohort were invited to participate in a COVID-19 impact survey in May-June 2020 (n=1321). Parents were asked about the impact of the pandemic and their intention to vaccinate their child against COVID-19 should a vaccine be approved. Past infant vaccination status was validated against public health records. Multinomial regression models were run to estimate associations between demographic factors, past vaccination status, and vaccine intention. Qualitative responses regarding factors impacting decision making were analyzed thematically.

**Results:** Sixty percent of parents (n=798) intended to vaccinate their children, but 9% (n=113) said they did not intend to vaccinate and 31% (n=410) were unsure. Lower education and income were inversely associated with intention to vaccinate. Incomplete vaccination history was associated with intention not to vaccinate but not uncertainty. Qualitative responses revealed concerns over vaccine safety and efficacy, long term effects and a rushed vaccination process.

**Interpretation:** Almost a third of parents remain unsure about vaccinating their children against COVID-19, even within a group with historically high uptake of infant vaccines. Given the many uncertainties about future COVID-19 vaccines, clear communication regarding safety will be critical to ensuring vaccine uptake.

## Introduction

In Canada, as of November 2020, there have been more than 300,000 people infected with severe acute respiratory syndrome coronavirus 2 (SARS-CoV-2) and over 10,000 deaths due to Coronavirus disease 2019 (COVID-19), with over 1 million deaths worldwide.^1, 2^ Given the devastating human, economic and social cost of the pandemic, the development of a vaccine remains a critical strategy to mitigate its impact.^3^ However, the development of a vaccine is not sufficient, as modeling suggests that up to 80% of the population needs to receive a vaccine that is 70% effective in order to end the pandemic without additional non-pharmaceutical interventions (e.g. physical distancing, masks, etc).^4^ Vaccine uptake relies on adequate production and distribution, but also on high levels of vaccine acceptance among the general public.^5^

Emerging studies with adults suggest that 60-80% are willing to receive a COVID-19 vaccine and 10% are not, with the remaining being unsure.^5-8^ Older age, higher education and higher income are associated with increased willingness to be vaccinated for COVID-19.^5-7^ In Canada, the National Advisory Committee on Immunization (NACI) has identified prioritized groups for early COVID-19 immunization.^9^ While NACI doesn’t currently identify children as a priority population unless they have other underlying risk factors, they will be eligible for vaccination once sufficient vaccine supply is available. If the evolving evidence finds that children are important transmitters of infection to more vulnerable populations, and if COVID-19 disease transmission within schools continues to grow, vaccination of children will become increasingly important. Understanding what factors impact parental decision-making prior to vaccine roll-out is critical for early engagement with parents about COVID-19 vaccine intentions and ensuring adequate uptake for COVID-19 infection control. A limited number of cross-sectional studies, primarily outside of Canada, have asked if parents would be willing to vaccinate their children, with acceptance ranging from 65% to 75%, but motivations for not vaccinating remain understudied.^10-12^ Past practices around vaccination may be critical to understanding barriers to uptake. Using longitudinally collected data provides the most accurate description of past vaccination behaviour. Moreover, understanding what factors influence decision-making will be critical to understanding how to communicate about a potential new vaccine. Thus, the objectives of this study are to understand parents’ COVID-19 vaccine intentions, using longitudinal data including historically collected vaccination information and demographic factors, and to explore reasons for and against COVID-19 vaccination.

## Methods

### Participants

This study used data from the longitudinal cohort study All Our Families in Alberta, Canada. Cohort characteristics and study design are described in detail elsewhere.^13^ Briefly, the All Our Families Cohort is a population-based pregnancy cohort that began in 2008 and recruited over a three year period. Of the 3388 women originally enrolled, 2455 remain part of the study after 12 years (72%). From May-June 2020, All Our Families participants, whose children had reached ages 9-12 years, were invited to complete the COVID-19 impact survey. Of the 2455 eligible participants, 1321 responded (53.8%). This study received ethical approval from the Conjoint Research Ethics Board of the University of Calgary.

### Measures

The COVID-19 Impact survey asked a series of questions about COVID-19 infection, job loss, and the impact of school closures and physical isolation measures on mental health and social connections. Participants reported on vaccine intentions for their All Our Families child. Specifically, participants were asked: “If a COVID-19 vaccine is approved, would you vaccinate your child?” (no, yes, unsure). Participants had the opportunity to provide a narrative response on what would impact their decision to vaccinate using an open text box. Participant responses were linked to previously collected longitudinal data on child’s past infant vaccine status at age 2 and demographic characteristics. Vaccine coverage at age 2 was collected via parental report and validated against administrative public health records.^14^ Vaccine status at this age was categorized as “partially or not vaccination” or “complete vaccination”, according to the infant vaccine schedule in Alberta.

### Data Analysis

Descriptive statistics on demographic characteristics and responses to the COVID-19 impact survey of the sample are provided. To describe which families were least likely to vaccinate, or unsure whether to vaccinate, we estimated bivariate multinomial regression models to describe unadjusted associations between participant characteristics and intention to vaccinate their child against COVID-19. A complete case analysis was used due to low missing data (<1%). The reference category was ‘intend to vaccinate’. All quantitative analyses were carried out using SAS version 9.4. In order to understand the factors impacting vaccine intentions, the qualitative data from narrative responses were analysed. Responses were coded thematically, and then the coding scheme and analysis was validated in a random sample of 20% of responses by a second author. If participants cited more than one reason, their answers were coded to multiple categories.

## Results

### Quantitative results

Participant characteristics stratified by vaccine intention are shown in Table 1.

**Table 1:** Characteristics of parents’ their intention to have their child receive the COVID19 vaccine

|  | **Overall**<br>**(N = 1,321)**<br>**n (%)<sup>a</sup>** | **Yes**<br>**(N=798)**<br>**n (%)** | **No**<br>**(N=113)**<br>**n (%)** | **Unsure**<br>**(N = 410)**<br>**n (%)** |
| --- | --- | --- | --- | --- |
| Maternal Age (mean, sd) – range 28-57 | 42.2 (4.4) | 42.5 (4.1) | 41.9 (4.7) | 41.5 (4.7) |
| Maternal Education (high school or less)<sup>b</sup> | 236 (17.9) | 106 (13.3) | 34 (30.1) | 96 (23.5) |
| Family Income (<\$80,000)<sup>a, c</sup> | 198 (15.1) | 97 (12.8) | 29 (26.1) | 72 (17.7) |
| Marital Status (Single/Divorced/Separated/Widowed)<sup>c, d</sup> | 69 (5.5) | 42 (5.5) | 7 (6.4) | 20 (5.1) |
| Ethnicity (self-identified minority)<sup>e</sup> | 221 (16.8) | 132 (16.6) | 20 (19.9) | 69 (17.0) |
| Child vaccine history (partial/not vaccinated)<sup>f</sup> | 200 (15.1) | 102 (12.8) | 33 (29.2) | 65 (15.9) |
| COVID19 Infection (Yes/Maybe)<sup>g</sup> | 80 (6.1) | 54 (6.8) | 7 (6.2) | 19 (4.7) |
| a) slight variation in the denominator due to missing data on income or marital status (<1%); b) ref: some post secondary/completed college/undergraduate or higher; c) ref: family income (≥\$80,000); d) ref: married/common-law; e) ref: self-identified White; f) ref: complete vaccines at 2 years; g) ref: Not infected | | | | |

Approximately 60% (n=798) of participants reported that they intended to give their 9-12 year old child the COVID-19 vaccine, 9% (n=113) would not, and 31% (n=410) were unsure. The mean age of mothers was 42 years, and 82% had a completed post-secondary degree or higher. Fifteen percent of children of participants had partial or no vaccinations at age 2 (12.0% and 3.2%, respectively). Only 1.1% of families had a confirmed COVID-19 infection at the time of the survey, while another 5.0% had a suspected case. Multinomial analyses linking factors to vaccine intention status are shown in Table 2 with statistically significant results (p<0.05 in bold).

**Table 2:** Odd Ratios (OR) from multinomial models for parents reporting their intention to have their child receive the COVID19 vaccine.

|  | No vs. Yes<br>OR (95% CI) | Unsure vs. Yes<br>OR (95% CI) |
| --- | --- | --- |
| Maternal Age (mean, sd) <sup>a</sup> | 0.97 (0.93, 1.01) | **0.94 (0.92, 0.97)** |
| Maternal Education (high school or less) <sup>b</sup> | **2.80 (1.78, 4.40)** | **1.98 (1.47, 2.71)** |
| Family Income (<\$80,000) <sup>c</sup> | **2.53 (1.58, 4.06)** | **1.53 (1.10, 2.14)** |
| Marital Status <sup>d</sup> (Single/Divorced/Separated/Widowed) | 1.16 (0.51, 2.66) | 0.91 (0.53, 1.58) |
| Ethnicity (self-identified minority) <sup>e</sup> | 1.09 (0.65, 1.83) | 1.02 (0.74, 1.41) |
| Child vaccine history (partial/not vaccinated) <sup>f</sup> | **2.81 (1.78, 4.40)** | 1.29 (0.92, 1.80) |
| COVID19 Infection (Yes/Maybe) <sup>g</sup> | 0.91 (0.40, 2.05) | 0.67 (0.39, 1.15) |
| All ORs represent bivariate associations and are unadjusted for other factors. a) increasing in years; b) ref: completed college/undergraduate or higher; c) ref: family income ($\geq \$80,000$); d) ref: married/common-law; e) ref: self-identified White; f) ref: complete vaccines at 2 years; g) ref: Not infected | | |

Lower socio-economic status was associated with vaccine intention, with a stronger association for those who do not intend to vaccinate, compared to those who are unsure. Participants with less education were more likely to not want to vaccinate (OR 2.80, 95% CI 1.78, 4.40) or be unsure (OR: 1.98, 95%CI 1.47, 2.71). A similar pattern was seen for income. History of partial or non-vaccination was associated with intent to not to vaccinate (OR 2.81, 95%CI:1.78, 4.40). There was no association between vaccination history and uncertainty regarding a COVID-19 vaccine (OR 1.29, 95%CI: 0.92, 1.80).

### Qualitative results

Eighty-five percent of participants provided a response in the narrative text box asking about reasons underlying vaccine intention. Thematic analysis revealed ten primary factors influencing decision-making were identified among all parents, regardless of intention to vaccinate. Percent of respondents in each category (yes, no, unsure) listing a specific factor and are presented in Figure 1. The inter-rater agreement for the categorization of narrative responses into themes was 82% (kappa 0.76).

**Figure 1:**
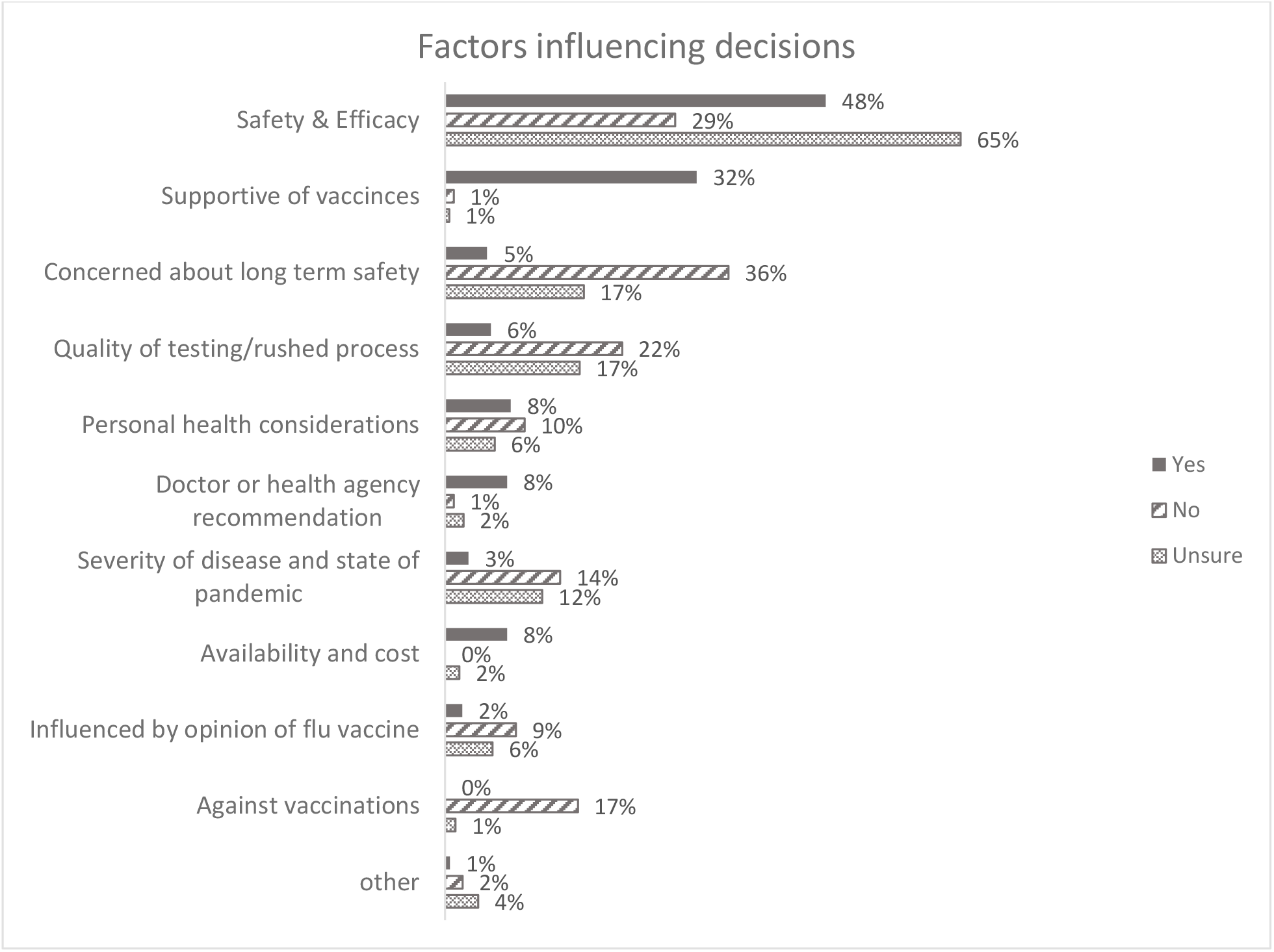
Factors influencing vaccine intentions as thematically coded from narrative responses. Percentages are within each intention category (i.e. 48% of those intending to vaccinate mentioned the first theme, safety and efficacy). Categories are not mutually exclusive and may add up to more than 100%

A description of each theme, and example quotes are provided in Table 3.

**Table 3:** Quotes for qualitative categories.

| Factor | Quotes |
| --- | --- |
| Safety and efficacy | "The effectiveness of the vaccine is key to consider and any side effects."<br>"I believe in vaccinations so would lead toward vaccinating - but would need more scientific information before making the final decision." |
| Supportive of vaccines | "would 100% vaccinate my family as soon as possible, we 100% support vaccinations."<br>"I believe in vaccines, I believe in science." |
| Long term safety | "Trial period will be too short to predict all possible long-term risks. if a few years, maybe consider, definitely not within 1-2 years."<br>"I am hesitant to take a vaccine or have my child injected with a vaccine that is so new. I would be afraid of complications in futures years that are now unknown." |
| Rushed process /Scientific quality | "It's safety. It seems like this vaccine is being rushed through trials."<br>"I trust medicine and science and have always vaccinated in the past; my only hesitancy with this vaccine would be the 'desperation/rush' that everyone is looking for a 'cure/solution' to COVID" |
| Perception of personal risk | "I am immune compromised so the family will be getting it to protect my health"<br>"If we have any underlying health issues that would compromise our immune system."<br>"I will not be receiving vaccine because I feel it's useless to us, we are strong enough to get over this flue," |
| Recommendation from doctor or health authority | "The recommendation of the Public Health Agency of Canada ."<br>"My doctor's recommendation would be the only opinion I would use to make my decision about being vaccinated." |
| Perception of risk from COVID-19 | "Risk of contracting Covid-19 would need to be greater than any risk associated to the vaccine."<br>"How much of the virus is still going on and impacting society." |
| Availability and cost | "Availability. I would pay for it if it was reasonable and available in my city."<br>"Availability and ease of access - I would not want to be standing in huge long lines for hours waiting for the vaccine like I did with H1N1." |
| Attitude towards flu vaccine (both positive and negative) | "My child has had all of the childhood vaccines but our family does not obtain the influenza vaccine as I feel we are all very healthy."<br>"Like the flu shot, is it really going to get the right strand of COVID-19?"<br>"We get the flu shot each year to protect our family from the worst of the effects of the flu." |
| Against vaccinations | "I would not get the vaccine or give it to my children. If there were any measures to make the vaccine mandatory or if people with the vaccine were given preferential treatment it would further solidify my stance to not get the vaccine. The other factor that would affect my decision is the overbearing influence on WHO, Health Canada/PHAC and AHS from corporate entities."<br>"Nothing will impact or change my view to vaccinate. I will not vaccine anyone in my family" |
| Other (mandatory, family opinions) | "If it is mandatory for work and school"<br>"My ex husband is not for vaccines. This will be my challenge." |

The most common factor mentioned was “safety and efficacy”, which included concerns about potential side-effects of vaccination. However, those intending not to vaccinate were more likely to mention long term safety (36%) than general safety and efficacy (29%). In addition, concerns regarding the rushed nature of testing which could potentially compromise the safety of the vaccine was cited among all groups (yes: 6%, no: 22% and unsure: 17%). Personal health conditions were noted among those intending to vaccinate (8%) those not intending to vaccinate (10%) and those who were unsure (6%). Four percent of participants overall mentioned their attitude toward the influenza vaccine impacting their thoughts on a COVID-19 vaccine but to differing degree (2% among yes, 9% among no, and 6% among unsure). For example, some who were unsure said they thought the influenza vaccine was ineffective, however, some intending to vaccinate said they got their flu shot every year and would also get a COVID-19 vaccine. Among those not intending to vaccinate, or unsure, belief that the COVID-19 disease was not very severe, or that the pandemic would be over soon, were also factors cited in their decision making.

### Interpretation

Among families for with generally high levels of complete infant vaccinations, 31% reported they were unsure and 9% reported they would not vaccinate their child against COVID-19. Findings from both the quantitative and qualitative analysis suggest three key messages outlined in detail below. First, incomplete infant vaccination was associated with negative intentions towards a COVID-19 vaccine, but not uncertainty. Second, attitudes towards a COVID-19 vaccine may reflect growing uncertainty about vaccine testing and development. Third, clear communication around the COVID-19 vaccine will be critical to assuage fears about a novel vaccine. While children may not be among the first to receive COVID-19 vaccinations in Canada, understanding parental motivations remains critical for ensuring high uptake once a vaccine is rolled out for this age group.^9^

Families characterized by less education and income were more likely to have negative or uncertain intentions towards vaccinating their child against COVID-19, consistent with studies on COVID-19 vaccine intentions in adults.^6-8^ A history of partial or incomplete infant vaccination, was associated with not wanting to vaccinate against COVID-19. Our findings are consistent with a multi-country study which found that having their child up to date on childhood vaccines was associated with COVID-19 vaccine acceptance in parents.^12^ Our study expands this knowledge by showing that having complete infant vaccinations was not associated with COVID-19 vaccine uncertainty. Interestingly, approximately 4% of participants mentioned that their thinking about a COVID-19 vaccine was influenced by their attitude toward the influenza vaccine, with both positive and negative opinions. This suggests that attitudes towards the influenza vaccine are more salient than attitudes toward childhood vaccines when it comes to COVID-19 vaccine intentions. The influenza vaccine has historically had lower uptake than childhood vaccines.^15^ Given the very high proportion of parents who remain uncertain about a COVID-19 vaccine, reliance on parental attitudes toward childhood vaccinations may not be sufficient for broad uptake of a COVID-19 vaccine.

The qualitative data showed that the majority of families had concerns around safety and efficacy of the vaccine, which has been noted previously.^10^ However, responses from “no” and “unsure” participants specifically mentioned concerns around long term safety. Among those not intending to vaccinate, 36% cited long term safety compared to 29% noting general safety concerns. Respondents noted the need for years of testing or a guarantee of 100% safety which may reflect unrealistic expectations for vaccines.^16, 17^ Polarization regarding vaccines is increasingly common, and has been linked with political ideology and general skepticism of science in both Europe and the United States.^6, 18, 19^ In Canada, vaccine hesitancy has increased in recent years and careful engagement with those who may be uncertain about vaccines is recommended.^20^ Moreover, only 1% of “no’s” and 2% of “unsure’s” mentioned willingness to rely on the recommendation from a doctor or public health authority, compared to 8% of “yeses”. Longer term engagement with non-combative strategies involving health care providers and public health leaders may be critical for reengaging those who remain skeptical about vaccines.^20^

Finally, among those who did not intend to vaccinate, or were unsure, a considerable number cited mistrust or concern with the rushed nature of testing. With increasing focus on novel vaccine types and preliminary promising findings with novel types of vaccines, there is a need to effectively communicate about the development, safety and efficacy of these vaccines.^16, 21^ And while these new developments may hold promise, the consequences on overall vaccine confidence could be severely threatened if novel vaccines have unintended consequences.^22, 23^ Due to the scope of the pandemic, vaccine trials are increasingly being highlighted in mainstream media and data from Canada and Australia suggests that acceptance of a new COVID-19 vaccine is declining over time.^24, 25^ Clear communication around risks and benefits will be critical and research into effective communication strategies around novel vaccines is urgently needed.^16, 26^

This study may have limited generalizability due to participant characteristics and response rate (54%). Responders were generally more affluent and more educated than non-respondents, and our sample reflected a population with generally higher complete infant vaccination than the average in Alberta or other Canadian provinces.^27, 28^ This would likely underestimate attitudes against a COVID-19 vaccine and could bias associations towards the null. Moreover, data were collected in the first wave of COVID-19 (May-June 2020) and attitudes toward vaccine intentions may change over the course of the pandemic. We only had validated information for vaccination status up to age 2. While 85% of participants indicated that their child’s vaccines were complete by age 2, this might have dropped off if we had complete data on pre-school vaccination status. We also did not have recent information on flu vaccine uptake, which has been associated with COVID-19 vaccine studies in adults.^7^ Follow-up research will more carefully assess influenza vaccine attitudes as well as the how parents view the risks of COVID-19 infection compared to a novel COVID-19 vaccine.

Families with lower income or lower education may be more reluctant to accept a COVID-19 vaccine. Complete infant vaccination was not associated with uncertainty about a COVID-19 vaccine, suggesting parents may view a novel COVID-19 vaccine differently from traditional infant immunizations. Moreover, our population had higher average rates of complete infant vaccination, suggesting that positive COVID-19 vaccination intentions may be even lower than our reported 60%. In order to maximize children’s uptake of a COVID-19 vaccine, assuaging parents’ concerns regarding safety, efficacy and testing appears to be paramount. Targeted public health strategies that include clear communication about safety and efficacy, may increase acceptance. Emphasis on quality of scientific evidence may be particularly salient among parents who are unsure.

## Supporting information

reporting checklist

## Data Availability

Individual, de-identified data are available to researchers who provide a methodologically sound proposal for research. Inquires regarding access to data dictionaries, study protocols and questionnaires may be made at: https://allourfamiliesstudy.com/data-access/

## Acknowledgments

The authors acknowledge the All Our Families research team and thank the participants who took part in the study.

